# A cross-sectional study on the drug usage of insomnia in the Chinese medicine hospital in Long gang, Shenzhen

**DOI:** 10.1101/2020.12.28.20248931

**Authors:** Jingfeng Lin, Zhenyi Wang, Danfeng Tian, Run Xi, Zhenyun Han

**Affiliations:** Beijing University of Chinese Medicine, Beijing, 100029, China; Shenzhen Hospital of Beijing University of Chinese Medicine (Longgang), Shenzhen, 518100, China

**Keywords:** Shenzhen, insomnia, drug usage

## Abstract

**Introduction:** Insomnia was widely distributed among the population, and it was a risk factor for many diseases. To evaluate the condition of usage of drugs in Chinese hospital of Longgang, Shenzhen, we carried this cross-sectional research. We extracted the information of drug usage, symptoms of patients by R software (version 4.0.2) from Hospital Information System (HIS). The research was registered in Chinese Clinical Trial Registry, ChiCTR2000040703.

**Methods:** A retrospective, cross-sectional study was conducted in Shenzhen Hospital of Beijing University of Chinese Medicine (Long Gang). Insomnia patients from Jan 1, 2016 to Nov 10, 2020 were included to cross-sectional study. We analyzed the basic information, the condition of drug usage and the relation of symptoms and drug usage by R software (version 4.0.2).

**Results:** Totally 9439 patients were included in the study. The average age of these patients was 45.81years (SD 13.97 years). Anxiety, dreaminess, dizzy, palpitation, headache, thirsty, weakness, chest distress, annoyance, abdominal distension, bad moods, difficulty falling asleep and bitter taste were core symptoms of insomnia. Totally 14256 times (67.65%) patients received drug prescription and more than one insomnia drug was administered in 8355 patients. The 10 most used drugs ranged from more to less were Estazolam(29.99%), Zaoren Anshen Capsule(15.50%), Oryzanol(14.82%), Diazepam(14.51%), Flupentixol and Melitracen(14.30%), Alprazolam(8.12%), Zolpidem Tartrate(5.29%), vitamin B6(4.76%), Sertraline(4.03%), Clonazepam(2.97%).

**Conclusion:** The drug usage for insomnia in the Chinese medicine hospital in Long gang, Shenzhen were mainly included benzodiazepine, nonbenzodiazepine, Chinese patent medicines, anti-anxiety and anti-depression drugs, oryzanol and vitamin B6. The usage of Oryzanol and vitamin B6 should be abused in Chinese medicine hospital, and the usage of Chinese medicine should be more rigorous evaluated. The nonbenzodiazepine should be promoted and broader understood in Chinese medicine hospital in Longgang, Shenzhen.

## 1. Introduction

Insomnia is broadly defined as dissatisfaction with sleep either qualitatively or quantitatively^1^. The rates of insomnia varied from 6%-19%^2 3^. Classical benzodiazepines and benzodiazepine receptor agonists (BZRAs, Nonbenzodiazepines; zopiclone, zolpidem, and zaleplon) are used in managing insomnia, but antidepressants are also used in clinical practice, although there is less evidence of their efficacy^4^ In additional, antipsychotics, antihistamines, phytotherapeutic substances and melatonin were also used as the therapeutic regimen for insomnia with lower evidence^2^. Insomnia could be a risk factor for cardiovascular diseases^5-7^, arterial hypertension, myocardial infarction, chronic heart failure^8 9^, type 2 diabetes^10^ and obesity and hypertension^11^. So, a big data analysis on the drug usage of insomnia was very necessary to help evaluating the reasonable of it. In China, to evaluate the condition of usage of drugs in Longgang Shenzhen, we carried this cross-sectional research. We extracted the information of drug usage, symptoms of patients by R software (version 4.0.2) from Hospital Information System (HIS). The association rules analysis was carried to find out the relationship with symptoms and usage of drugs. The research was registered in Chinese Clinical Trial Registry, ChiCTR2000040703.

## 2. Methods

### 2.1 Study setting and patients

A retrospective, cross-sectional study was conducted in Shenzhen Hospital of Beijing University of Chinese Medicine(Long Gang). Insomnia patients from Jan 1, 2016 to Nov 10, 2020 were included to cross-sectional study. The included criteria were: (1) International Classification of Sleep Disorders (ISCD-3) Diagnostic Criteria for short-term and chronic insomnia. (2) We excluded patients that were of mental or nervous system dysfunction, or unable to express willingness. This study was approved by the ethics committee of Shenzhen Hospital of Beijing University of Chinese Medicine(Long Gang).

### 2.2 Data collection and analysis

The information of patient, medication, and microbiologic examination, including register number, age, sex, diagnosis, admission time were collected from Shenzhen Hospital of Beijing University of Chinese Medicine Hospital Information System (HIS) by clinical practitioners. Data were analyzed by R software(version 4.0.2).

## 3. Results

### 3.1 Baseline patient characteristics

Totally 9439 patients were included in the study, with 21073 times coming to the doctors, among who 3470 (39.6%) were male, and 5699 (60.4%) were female. With there were 6577 patients (69.7%) and 14256(67.7%) times coming included in the retrospective cross-sectional study to assess the drug use. The average age of these patients was 45.81years (SD 13.97 years). Table 1 shows the characteristics of 9439 included patients.

**Table 1:** The characteristics of 9439 included patients.

|  | All | Male | female | P-value |
| --- | --- | --- | --- | --- |
| **Average age** |  |  |  |  |
| Mean (SD) | 45.81 (±13.97) | 45.28 (±13.92) | 46.16 (±14.00) | 0.001 |
| **Agegroup** |  |  |  |  |
| ≤30 | 1154 (12.23%) | 456 (39.51%) | 698 (60.49%) | 0.0005 |
| 31-45 | 3740 (39.63%) | 1573 (42.06%) | 2167 (57.94%) |  |
| 46-60 | 3050 (32.32%) | 1164 (38.16%) | 1886 (61.84%) |  |
| >60 | 1493 (15.82%) | 547 (36.64%) | 946 (63.36%) |  |

\*Excluded the patients with no information of age

### 3.2 Use of benzodiazepine

Among the total of 21073 times, 14256 times (67.65%) patients received drug prescription and more than one insomnia drug was administered in 8355 patients. Among the patients using with benzodiazepines, 4276(29.99%) patients received estazolam, 1157(8.12%) patients received Alprazolam, 2069(14.51%) patients received Diazepam,. Among the 754(5.29%) patients received Zolpidem Tartrate, 424(2.97%) patients received Clonazepam.

### 3.3 Use of nonbenzodiazepine

Dexzopiclone 119 0.83%

Among the patients using with nonbenzodiazepines, 754(5.29%) patients received Zolpidem Tartrate, 119(0.83%) patients received Dexzopiclone.

### 3.4 Use of Chinese patent medicines

Totally 2209(15.50%) patients received Zaoren Anshen Capsule and 400 (2.81%) patients received Shumian Capsule.

### 3.5 Use of traditional Chinese medicines

Among 4406(30.91%) comings used traditional Chinese medicines.

### 3.6 Use of anti-anxiety and anti-depressants

Among the all patients, 2038 (14.3%) were given Flupentixol and Melitracen. And of the 575(4.03%) patients took Sertraline.

### 3.7 Use of other kinds of medicine

Among the all patients, 2113 (14.82%) were given Oryzanol. And of the 679(4.76%) patients took Vitamin B6.

### 3.8 Symptoms collection

A total of 17,337 outpatient electronic medical records (EMRs) were obtained by screening the included patient, and the retention of EMRs accounted for 82.27% of the total number of patients. The R software stringr package was used to extract the symptoms and signs information contained in the medical records. The extracted symptom and sign information results were sorted in descending order. The top 20 symptoms were selected, and the results were as follows:

From Table 7, we could conclude that anxiety, dreaminess, dizzy, palpitation, headache, thirsty, weakness, chest distress, annoyance, abdominal distension, bad moods, difficulty falling asleep and bitter taste were core syndromes of insomnia.

**Table 2:** Use of benzodiazepine

| Drugs | Frequency | Rates |
| --- | --- | --- |
| Estazolam | 4276 | 29.99% |
| Diazepam | 2069 | 14.51% |
| Alprazolam | 1157 | 8.12% |
| Clonazepam | 424 | 2.97% |

**Table 3:** Use of nonbenzodiazepine

| Drugs | Frequency | Rates |
| --- | --- | --- |
| Zolpidem Tartrate | 754 | 5.29% |
| Dexzopiclone | 119 | 0.83% |

**Table 4:** Use of Chinese patent medicines

| Drugs | Frequency | Rates |
| --- | --- | --- |
| Zaoren Anshen Capsule | 2209 | 15.50% |
| Shumian Capsule | 400 | 2.81% |

**Table 5:** Use of anti-anxiety and anti-depressants

| Drugs | Frequency | Rates |
| --- | --- | --- |
| Flupentixol and Melitracen | 2038 | 14.30% |
| Sertraline | 575 | 4.03% |
| Escitalopram | 120 | 0.84% |

**Table 6:** Other kinds of medicine

| Drugs | Frequency | Rates |
| --- | --- | --- |
| Oryzanol | 2113 | 14.82% |
| vitamin B6 | 679 | 4.76% |

**Table 7:**
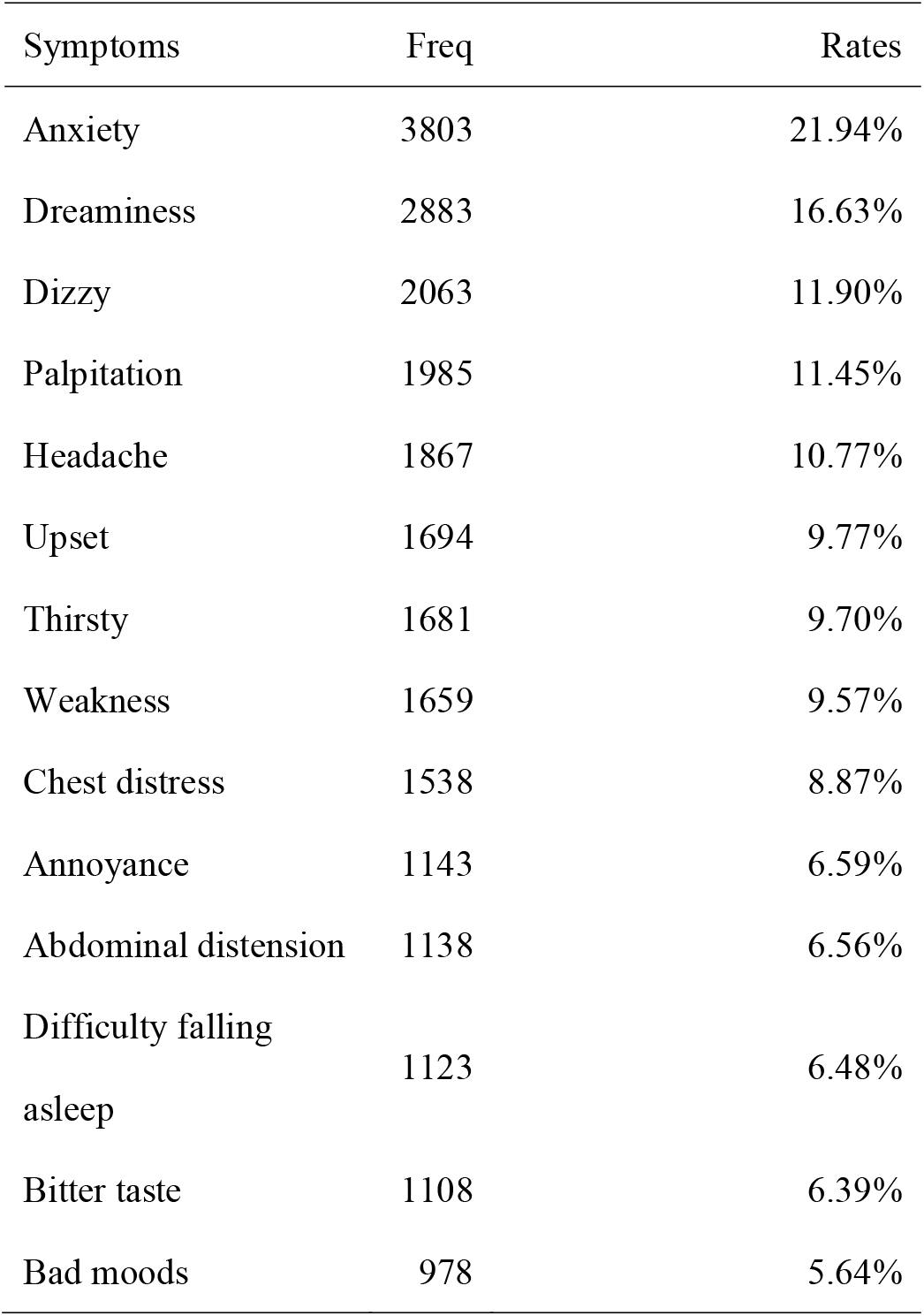
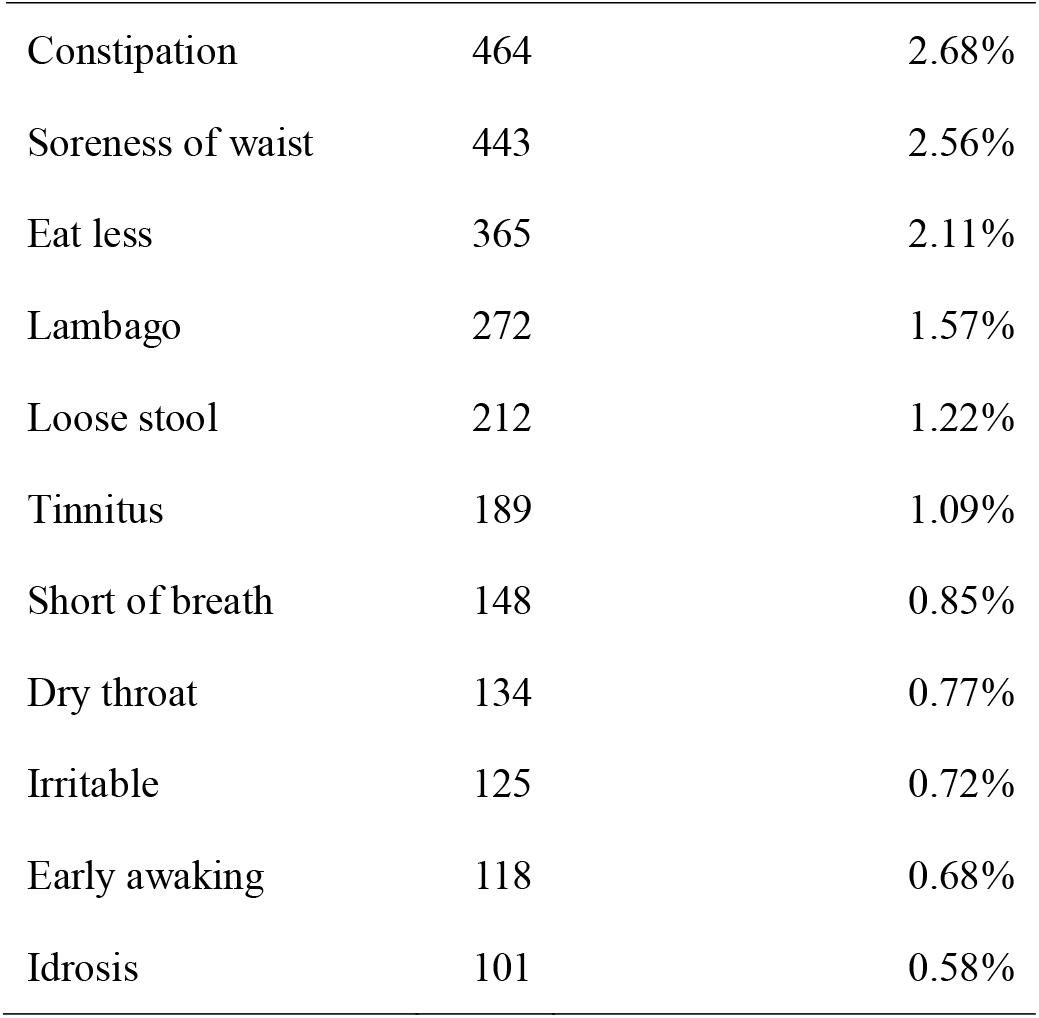
Top 20 symptoms of insomnia

| Symptoms | Freq | Rates |
| --- | --- | --- |
| Anxiety | 3803 | 21.94% |
| Dreaminess | 2883 | 16.63% |
| Dizzy | 2063 | 11.90% |
| Palpitation | 1985 | 11.45% |
| Headache | 1867 | 10.77% |
| Upset | 1694 | 9.77% |
| Thirsty | 1681 | 9.70% |
| Weakness | 1659 | 9.57% |
| Chest distress | 1538 | 8.87% |
| Annoyance | 1143 | 6.59% |
| Abdominal distension | 1138 | 6.56% |
| Difficulty falling<br>asleep | 1123 | 6.48% |
| Bitter taste | 1108 | 6.39% |
| Bad moods | 978 | 5.64% |
| Constipation | 464 | 2.68% |
| Soreness of waist | 443 | 2.56% |
| Eat less | 365 | 2.11% |
| Lambago | 272 | 1.57% |
| Loose stool | 212 | 1.22% |
| Tinnitus | 189 | 1.09% |
| Short of breath | 148 | 0.85% |
| Dry throat | 134 | 0.77% |
| Irritable | 125 | 0.72% |
| Early awaking | 118 | 0.68% |
| Idrosis | 101 | 0.58% |

**Table 8:**
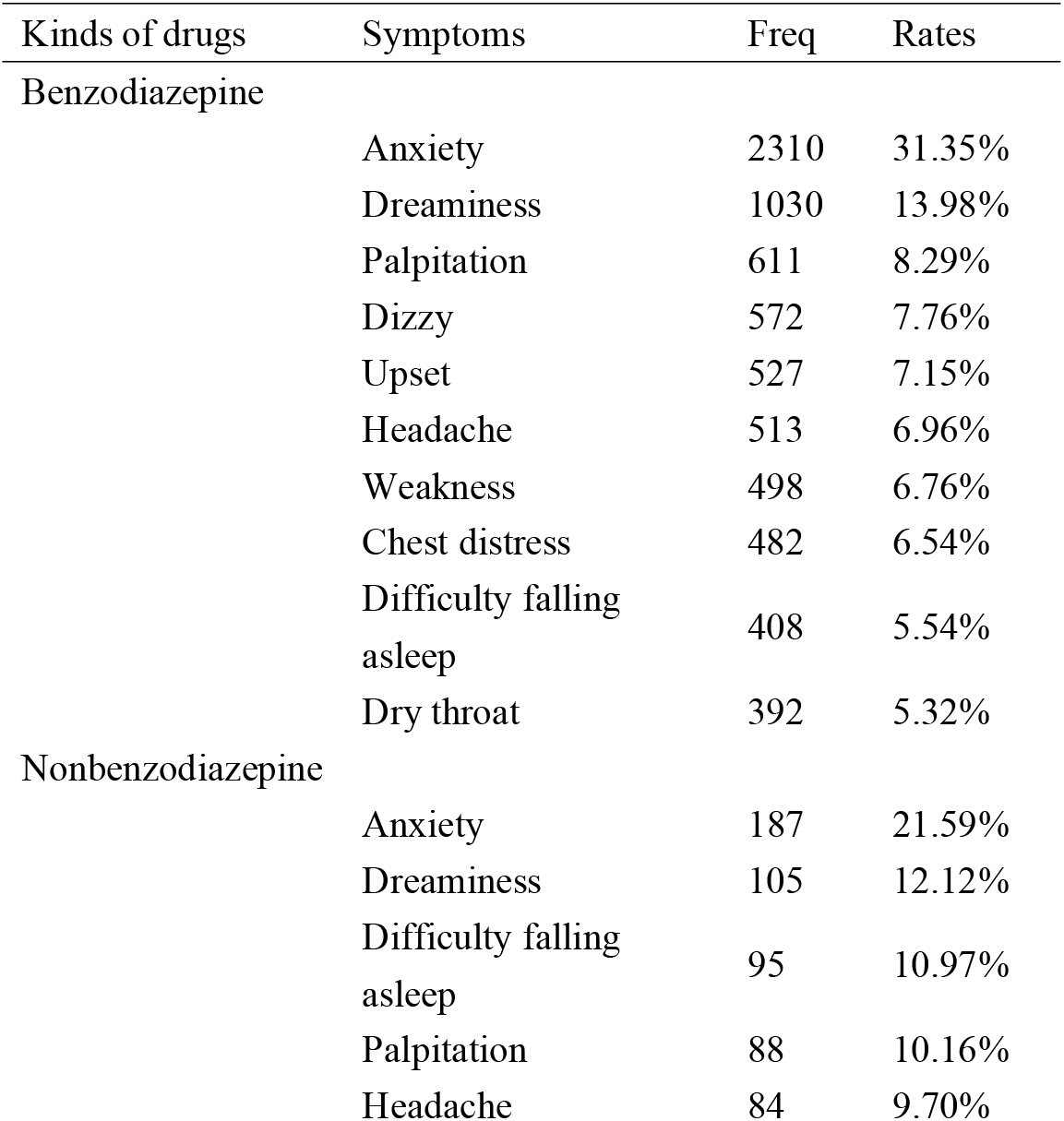
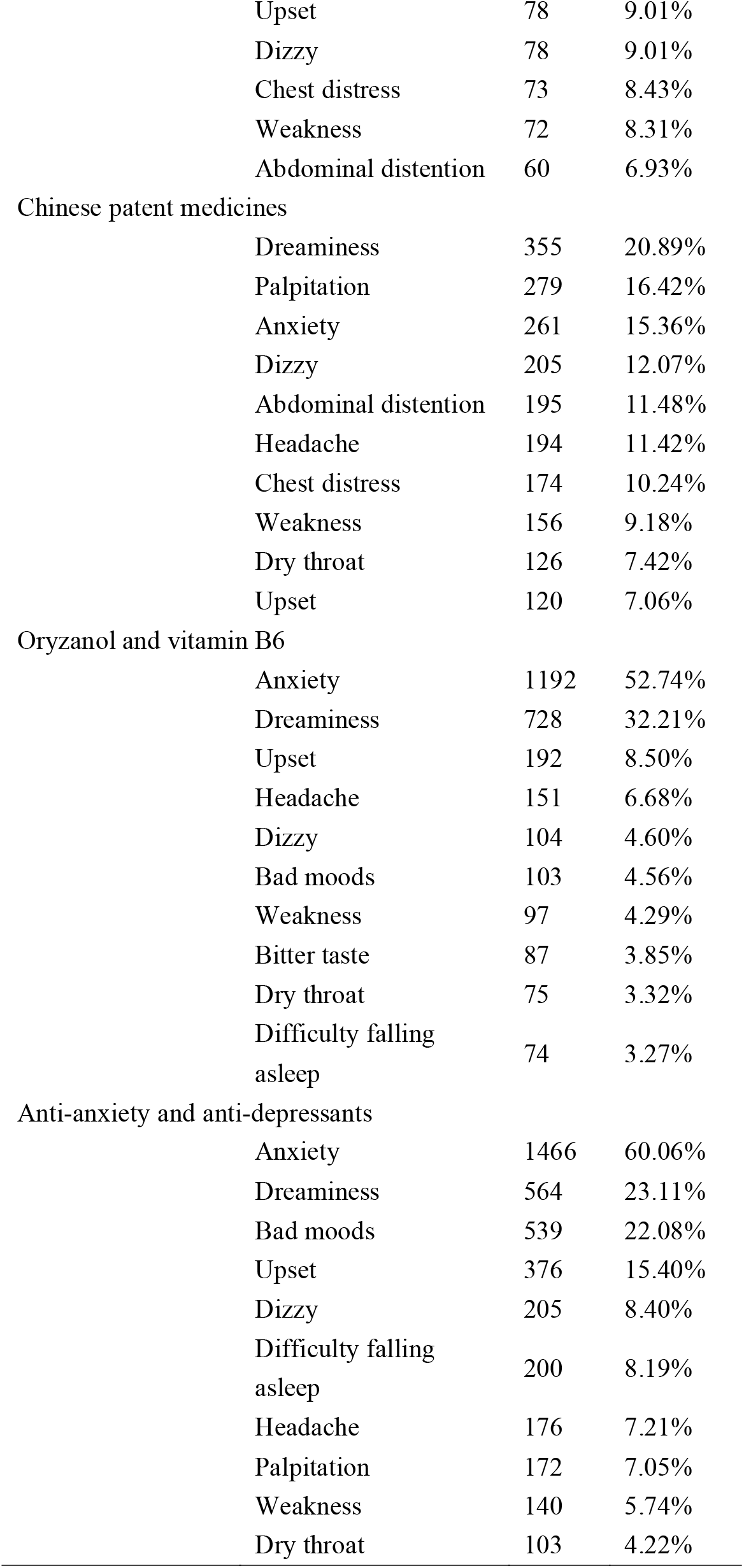
Different kinds of drugs and its main symptoms

### 3.9 Reinteraction of symptoms and drugs

We select the patients using benzodiazepine, nonbenzodiazepine, Chinese patent medicines, oryzanol and vitamin B6 respectively, anti-anxiety and anti-depressants and summary the times of syndromes appearing in the patients. The results were as following:

For the patients who received benzodiazepine, nonbenzodiazepine, Oryzanol and vitaminB6, and Anxiety and depression, most common symptom was Anxiety. However, for Chinese patent medicines, most common symptom was Dreaminess.

Association rules analysis was carried for various symptoms and different kinds of drugs by arules package, R software. In the analysis of association rules of kinds of drugs and symptoms, we selected association rules of support degree >0.01 and confidence degree >0.1 to find the association rules between benzodiazepine and symptoms, and we selected association rules of support degree >0.005 and confidence degree >0.05 to find the association rules between Chinese patent medicines, Oryzanol and Vitamin, Anti-anxiety and anti-depressants and symptoms. We also found that there were no significant association rules between Nonbenzodiazepines and symptoms. The aruleViz package was used to draw the correlation diagram, and the results were shown in Figure 1.

**Figure 1:**
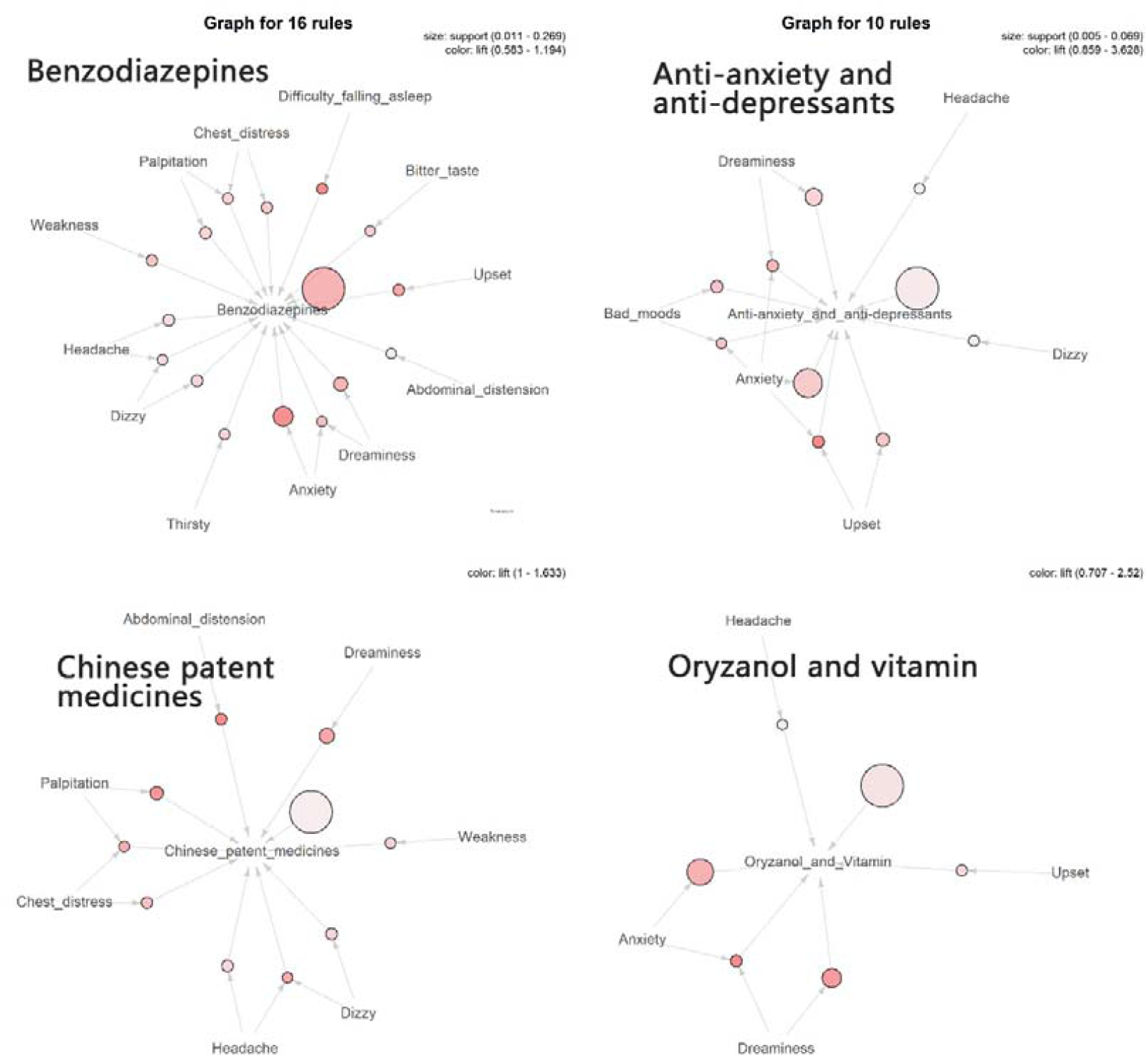
Association rules analysis of drug usage and symptoms.

The results showed that patients received benzodiazepines were associated with Anxiety, Dreaminess, Upset, Bitter taste, Difficulty falling asleep, Chest distress, Palpitation, Weakness, Headache, Dizzy, Thirsty, Abdominal distension, which were in accordance with the result in Table. Patients received Chinese patent medicines were associated with Dizzy, Weakness, Dreaminess, palpitation, chest distress and headache. Patients received Oryzanol and Vitamin were associated with Headache, anxiety, dreaminess and upset. Patients received anti-anxiety or anti-depression drugs were associated with anxiety, bad mood, upset, dizzy, dreaminess and headache.

## 4. Discussion

This study evaluates the drug use in treating the patients in Longgang, Shenzhen, who suffered from insomnia by Hospital Information System (HIS) in Shenzhen Hospital of Beijing University of Chinese Medicine(Long Gang). About 9439 patients were included in the study, with 21073 times coming to the doctors to assess the rationality of drug use and the majority of them are female(60.4%). The results demonstrate that benzodiazepine, nonbenzodiazepine, traditional Chinese medicines, Oryzanol and vitaminB6 are the principal drugs to treat insomnia in patients. Anti-anxiety and anti-depression are the main prescript drugs for insomnia with the symptoms of anxiety and depression.

In European insomnia guideline, the Benzodiazepines, benzodiazepine receptor agonists and some antidepressants are effective in the short term treatment of insomnia, and phytotherapeutics are not recommended for insomnia treatment^2^. In our research, we found that Chinese patent medicines(18.31%) and traditional Chinese medicines(30.91%) were widely used in Shenzhen Hospital of Beijing University of Chinese Medicine(Long Gang), which was contradicted to the guidelines. In other cross-sectional about drug usage for insomnia, Suzanne M. Bertisch^3^ did not reported the usage of phytotherapeutics in a big cross-sectional research of Prescription Medications for Insomnia in America. In past research, there were no significant evidence on the efficacy of phytotherapeutics for insomnia. Xiaojia Ni^12^ made a meta-analysis on efficacy of Chinese medicine on insomnia compared with placebo, and found that the Chinese medicine was better than placebo, however, the quality of included studies was pool^2^. So, we thought the efficacy of Chinese medicine on insomnia and the rationality of drug use in Shenzhen Hospital of Beijing University of Chinese Medicine(Long Gang) should be further evaluated.

For benzodiazepine and nonbenzodiazepine, a meta-analysis indicated that there were no difference between them^13^. In our research, we found that the usage of benzodiazepine was far more than nonbenzodiazepine, the reason we thought was the benzodiazepines were more acceptable in Longgang, Shenzhen doctors.

For Oryzanol and vitamin B6, there were about 14.82% patients received Oryzanol and about 4.76% patients received vitamin B6. In past research, there were no significant evidence indicated that Oryzanol could be helpful to insomnia, and it didn’t be recommended in guideline^13^. For vitamin B6, Patrick Lemoine^14^ found that the combination of melatonin, vitamin B6, and medicinal plants may be beneficial in mild-to-moderate insomnia. However, there were no single-evidence for vitamin B6 about whether it was effective for insomnia.

Alexander Winkler^15^ made an analysis on antidepression drugs and found that the efficacy of antidepressants was weaker than that for benzodiazepines and nonbenzodiazepines, other researches indicated that adverse effects and study withdrawals did not significantly differ between participants receiving doxepin and those receiving placebo^16^. In our research, Flupentixol and Melitracen and sertraline were mostly used in Beijing University of Chinese Medicine(Long Gang). For Flupentixol and Melitracen on insomnia, a research from China also reported the Flupentixol and Melitracen was widely used in 20 national hospitals of China, however, there was no precise evidence manifested the Flupentixol and Melitracen was effective to insomnia and was not be recommended in guideline^2^.

For sertraline, there were some evidence indicated that Antidepressant Pharmacotherapy could relief the symptoms in Patients With Comorbid Depression and Insomnia^17^ and the efficiency was exactly^2^.

For the relationship between symptoms and different kinds of drugs, we would find that antidepression and antianxiety drugs were related to Anxiety, bad moods and dreaminess, and Chinese patent medicines was more concretely about dreaminess, which indicated that the usage of antidepression and Chinese patent medicines was Symptom-related.

To improve the construction drug usage in Shenzhen, Longgang, we thought that the use of Chinese patent medicine and Chinese medicine should be evaluated more carefully. As it was well known that in Chinese medicine hospital in China, there were much Chinese medicine and Chinese patent medicine used than Western medicine hospital in China. However, the usage of Chinese medicine was lack of proper efficacy evaluation and clinical trial or observation with high level evidence^2^. The more rigorous, high-quality clinical trials should be carried to evaluate the exact efficacy in specific patients. In additional, we thought the nonbenzodiazepine had some advantages than benzodiazepine and should be promoted in Chinese medicine hospital.

For the deficiencies of this research, the data of research was from the HIS by R software, for some clinician would omit or miswrite some information by the busy work, the data could be some biases and the data maybe have some incompleteness. P. W. Handayani^18^ pointed that the HIS still needs to be refined in terms of providing optimal health services in development countries, and there was considerable dissatisfaction with the quality of the existing HIS among doctors. In addition, the information of patients in HIS of Chinese medicine hospital, Longgang, Shenzhen were mainly about sex, age, other important information was not recorded. So, as a sectional study, the information collection was inadequate.

In conclusion, the drug usage for insomnia in the Chinese medicine hospital in Long gang, Shenzhen were mainly included benzodiazepine, nonbenzodiazepine, Chinese patent medicines, anti-anxiety and anti-depression drugs, Oryzanol and vitaminB6. The usage of Oryzanol and vitaminB6 should be abused in Chinese medicine hospital, and the usage of Chinese medicine should be more rigorous evaluated. The nonbenzodiazepine should be promoted and broader understood in Chinese medicine hospital in Longgang, Shenzhen.

## Data Availability

The datasets used or analyzed during the current study are available from the corresponding author on reasonable request.

## Abbreviations

HIS: Hospital Information System
BZRA: Benzodiazepine receptor agonist

## Declarations

### Statement of Ethics

The experiment was approved by Ethic committee of Shenzhen Hospital of Beijing university of Chinese medicine.

### Authors contributions

Jingfeng Lin, Zhenyi Wang and Danfeng Tian performed the integral research. Jingfeng Lin and Run Xi performed data screen and data analysis. Zhenyun Han conceived and supervised the study. Jingfeng Lin, Zhenyi Wang and Danfeng Tian drafted the manuscript. All authors read and approved the final manuscript.

